# Harmonizing heterogeneous endpoints in COVID-19 trials without loss of information –an essential step to facilitate decision making

**DOI:** 10.1101/2020.03.31.20049007

**Authors:** Maja von Cube, Marlon Grodd, Martin Wolkewitz, Derek Hazard, Jerome Lambert

## Abstract

**Background:** Many trials are now underway to inform decision-makers on potential effects of treatments for COVID-19. To provide sufficient information for all involved decision-makers (clinicians, public health authorities, drug regulatory agencies) a multiplicity of endpoints must be considered. It is a challenge to generate detailed high quality evidence from data while ensuring fast availability and evaluation of the results.

**Methods:** We reviewed all interventional COVID-19 trials on Remdesivir, Lopinavir/ritonavir and Hydroxychloroquine registered in the National Library of Medicine (NLM) at the National Institutes of Health (NIH) and summarized the endpoints used to assess treatment effects. We propose a multistate model that harmonizes heterogeneous endpoints and differing lengths of follow-up within and between trials.

**Results:** There are currently, March 27, 2020, 23 registered interventional trials investigating the potential benefits of Remdesivir, Lopinavir/ritonavir and Hydroxychloroquine. The endpoints are highly heterogeneous. Follow-up for the primary endpoints ranges from four to 168 days.

A detailed precisely defined endpoint has been proposed by the global network REMAP-CAP, which is specialized on community-acquired pneumonia. Their seven-category endpoint accounts for major clinical events informative for all decision-makers. Moreover, the Core Outcome Measures in Effectiveness Trials (COMET) Initiative is currently working on a core outcome set. We propose a multistate model that accommodates analysis of these recommended endpoints. The model allows for a detailed investigation of treatment effects for various endpoints over the course of time thereby harmonizing differing endpoints and lengths of follow-up.

**Conclusion:** Multistate model analysis is a powerful tool to study clinically heterogeneous endpoints (mortality, discharge) as well as endpoints influencing hospital capacities (duration of hospitalization and ventilation) simultaneously over time. Our proposed model extracts all information available in the data and is - by harmonizing endpoints within and between trials - a step towards faster decision making. All ongoing clinical trials, especially those with severe cases, should accommodate primary analysis with a stacked probability plot of the major events mechanical ventilation, discharge alive and death.

## Introduction

Evidence for effective treatments to defeat COVID-19 is needed – fast. The number of registered randomized trials studying promising antiviral drugs is rising steadily. To ensure timely availability and evaluation of the results, it is essential that the studies are comparable. At the same time, the manifold ways treatment may have a beneficial effect need to be assessed and sufficient information for all decision makers must be provided (1,2). Thus, while a number of different endpoints is of major interest, comprehensive judgement is only possible if treatment effects on the various endpoints are presented simultaneously.

The Core Outcome Measures in Effectiveness Trials (COMET) Initiative is currently working on a core outcome set (COS) for COVID-19 randomized trials (3). In this manuscript, we propose a multistate model that harmonizes the multiplicity of endpoints while allowing for a detailed understanding of treatment effect (4,5). Thus, we do not aim at giving recommendation on suitable endpoints, but rather on additional statistical analysis that provides major insights for endpoints within, for example, the COS of COMET.

First, we provide a exemplarily overview of endpoints currently used in COVID-19 trials. To do so, we reviewed the primary endpoints of clinical trials registered in the database of the National Library of Medicine (NLM) at the National Institutes of Health (NIH) (6). We restricted the review to Remdesivir, Lopinavir/ritonavir and Hydroxychloroquine which are currently among the most promising treatment options for COVID-19. Then, we explain the multistate model approach. Even though the endpoints-review is restricted to a small number of treatments the proposed multistate methods apply equally to other COVID-19 clinical trials.

### Endpoints

We reviewed clinical COVID-19 trials to provide an overview of the primary endpoints used to analyze treatment effects. We included all randomized trials registered by March 27, 2020 in the clinical trial database of the NLM at the NIH with the primary purpose to study the efficacy of Remdesivir, Lopinavir/ritonavir or Hydroxychloroquine as treatment for COVID-19 diseased patients (6).

We found 23 trials that met the inclusion criteria. Of these trials, we extracted and categorized the definition of the primary endpoint. Moreover, we collected details on duration of follow-up, sample size and the patient population under study (Table 1 and Supplementary Table S1).

**Table 1:** Selected columns of Table S1 summarizing the results of the primary outcomes review. The category “unclear” for the columns mortality, mechanical ventilation and hospitalization indicates that we do not have sufficient information to judge whether this information is collected.

| **NCT Number** | **treatment** | **primary outcome** | **follow-up for primary outcome (days)** | **mortality** | **mechanical ventilation** | **hospitalization** |
| --- | --- | --- | --- | --- | --- | --- |
| NCT04292899 | Remdesivir | recovery | 14 | unclear | unclear | secondary |
| NCT04292730 | Remdesivir | discharge | 14 | unclear | unclear | primary |
| NCT04252664 | Remdesivir | recovery | 28 | secondary | secondary | unclear |
| NCT04257656 | Remdesivir | multistate (6-point ordinal scale) | 28 | primary | primary | primary |
| NCT04321616 | Remdesivir and Hydroxychloroquine | mortality | 21 | primary | secondary | secondary |
| NCT04280705 | Remdesivir | multistate (8-point ordinal scale) | 15 | primary | primary | primary |
| NCT04315948 | Remdesivir and Lopinavir/ritonavir and Hydroxychloroquine | multistate (7-point ordinal scale) | 15 | primary | primary | primary |
| NCT04307693 | Lopinavir/ritonavir and Hydroxychloroquine | viral load | 18 | secondary | secondary | unclear |
| NCT04286503 | Lopinavir/ritonavir | recovery | 30 | unclear | unclear | unclear |
| NCT04255017 | Lopinavir/ritonavir and Hydroxychloroquine | recovery | 14 | unclear | unclear | unclear |
| NCT04295551 | Lopinavir/ritonavir | recovery | 28 | unclear | unclear | unclear |
| NCT04251871 | Lopinavir/ritonavir | recovery | 28 | secondary | secondary | secondary |
| NCT04261907 | Lopinavir/ritonavir | clinical worsening | 14 | unclear | secondary | unclear |
| NCT04275388 | Lopinavir/ritonavir | recovery | 14 | unclear | unclear | unclear |
| NCT04276688 | Lopinavir/ritonavir | recovery | 30 | secondary | unclear | secondary |
| NCT04303299 | Lopinavir/ritonavir<br>and<br>Hydroxychloroquine | recovery | 168 | secondary | secondary | unclear |
| NCT04321993 | Lopinavir/ritonavir | multistate<br>(7-point<br>ordinal<br>scale) | 15 | primary | primary | primary |
| NCT04316377 | Hydroxychloroquine | viral load | 4 | secondary | secondary | secondary |
| NCT04323631 | Hydroxychloroquine | clinical<br>worsening | 28 | primary | unclear | unclear |
| NCT04315896 | Hydroxychloroquine | mortality | 120 | primary | secondary | secondary |
| NCT04322123 | Hydroxychloroquine | multistate<br>(6-point<br>ordinal<br>scale) | 15 | primary | primary | primary |
| NCT04321278 | Hydroxychloroquine | multistate<br>(7-point<br>ordinal<br>scale) | 15 | primary | primary | primary |
| NCT04261517 | Hydroxychloroquine | viral load<br>mortality | 14 | primary | unclear | unclear |

Five of the trials study Remdesivir, seven study Lopinavir/ritonavir and six study Hydroxychloroquine. Moreover, there are three trials studying both Lopinavir/ritonavir and Hydroxychloroquine, one studying Remdesivir and Hydroxychloroquine and one involving all of the three treatments.

The main part of the registered trials (n=9) uses a recovery outcome as primary endpoint. However, definition of recovery and duration of follow-up differ substantially between trials. Seven of these trials do not provide a precise clinical definition of their primary endpoint. Among those that provide more details, clinical recovery is defined in different ways. Follow-up in trials with recovery as endpoint ranges from 14 to 168 days with most trials (n=5) having a follow-up of 28 or 30 days.

Generally, follow-up ranges from four to 168 days. Most trials (n=16) have a follow-up of 14 to 15 days (n=6 and 5 respectively) or 28 days (n=5). Other outcomes are discharge (n=1), clinical worsening (n=2), mortality (n=3) and viral load (n=3; this includes one study with two primary endpoints).

When using a recovery endpoint, it is important to differentiate between community-acquired and hospital-acquired COVID-19 infection. Currently, cases in randomized trials are comprised of community-acquired infections. However, it cannot be excluded that in the near future hospital-acquired COVID-19 randomized trials will be necessary. Definition of recovery from COVID-19 in critically ill patients is complex and highly subjective due to the infeasibility to differentiating symptoms from COVID-19 and the initial illness (2).

The WHO R&D Blueprint expert group (7) recommends to use a categorical endpoint ranging from full recovery to death. For example, based on this recommendation Cao et al. (8) chose their primary endpoint as the time to clinical improvement, defined as the time from randomization to either an improvement of two points on a seven-category ordinal scale or discharge from the hospital, whichever came first. The seven-category ordinal scale of Cao et al. (8) consists of the following categories: 1, not hospitalized with resumption of normal activities; 2, not hospitalized, but unable to resume normal activities; 3, hospitalized, not requiring supplemental oxygen; 4, hospitalized, requiring supplemental oxygen; 5, hospitalized, requiring nasal high-flow oxygen therapy, noninvasive mechanical ventilation, or both; 6, hospitalized, requiring ECMO, invasive mechanical ventilation, or both; and 7, death. Six of the registered clinical trials use such a categorical endpoint. The number of categories varies between six and eight states.

Robust endpoints ensuring assessment of information for clinicians as well as public health authorities are duration of mechanical ventilation, hospitalization and mortality. Six (23%) studies consider mechanical ventilation in the primary endpoint and eight (35%) in the secondary endpoint. Similarly, hospitalization is included in the primary endpoint in seven (30%) of the studies and six (23%) in the secondary endpoint. Sixteen (70%) of the studies indicate that information on mortality is recorded (n=10, 43%) primary endpoint, (n=6, 23%, secondary endpoint). For other studies it is unclear whether this information is available. However, it can be presumed that more trials will record this information.

The heterogeneity in choice of endpoints reflects the manifold potential treatment effects, the various different decision makers involved and the different patient populations under study. Nonetheless, harmonization of the different endpoints is an essential step to fasten decision making of public health authorities and clinicians (2). At the same time, information on treatment effects should not be lost by oversimplification. Our proposal for harmonizing endpoints between and also within trials is based on information about major clinical events available in most of the registered clinical trials, namely mechanical ventilation, hospitalization and mortality.

### Statistical guidelines to analyze COVID-19 clinical trials

The categorical endpoint recommended by the WHO R&D Blueprint expert group (7) ensures detailed collection of information on a range of important events. Nonetheless, a single estimate for the treatment effect - albeit an important summary of the benefit on the primary outcome - does not provide sufficient information. All time-dynamic aspects of the treatment must be understood in detail to make well-informed decisions. In light of the emergency, it is essential to exploit the full potential of all the available data to be able to report on all potential treatment effects.

Multistate methodology is a powerful tool to study clinically opposite endpoints (mortality, discharge) as well as endpoints influencing hospital capacities (duration of hospitalization and ventilation) simultaneously over time (9,10). Duration of mechanical ventilation and hospitalization as well as mortality are robust endpoints providing important information for all decision makers.

To obtain a detailed understanding of potential treatment effects, we propose the multistate model shown in Figure 1. The boxes represent the possible states a patient may encounter and the arrows represent the possible transitions from one state to another. The model accounts for mechanical ventilation, discharge alive and death. Treatment effects on duration of mechanical ventilation, length of hospital stay and death are directly quantifiable.

**Figure 1:**
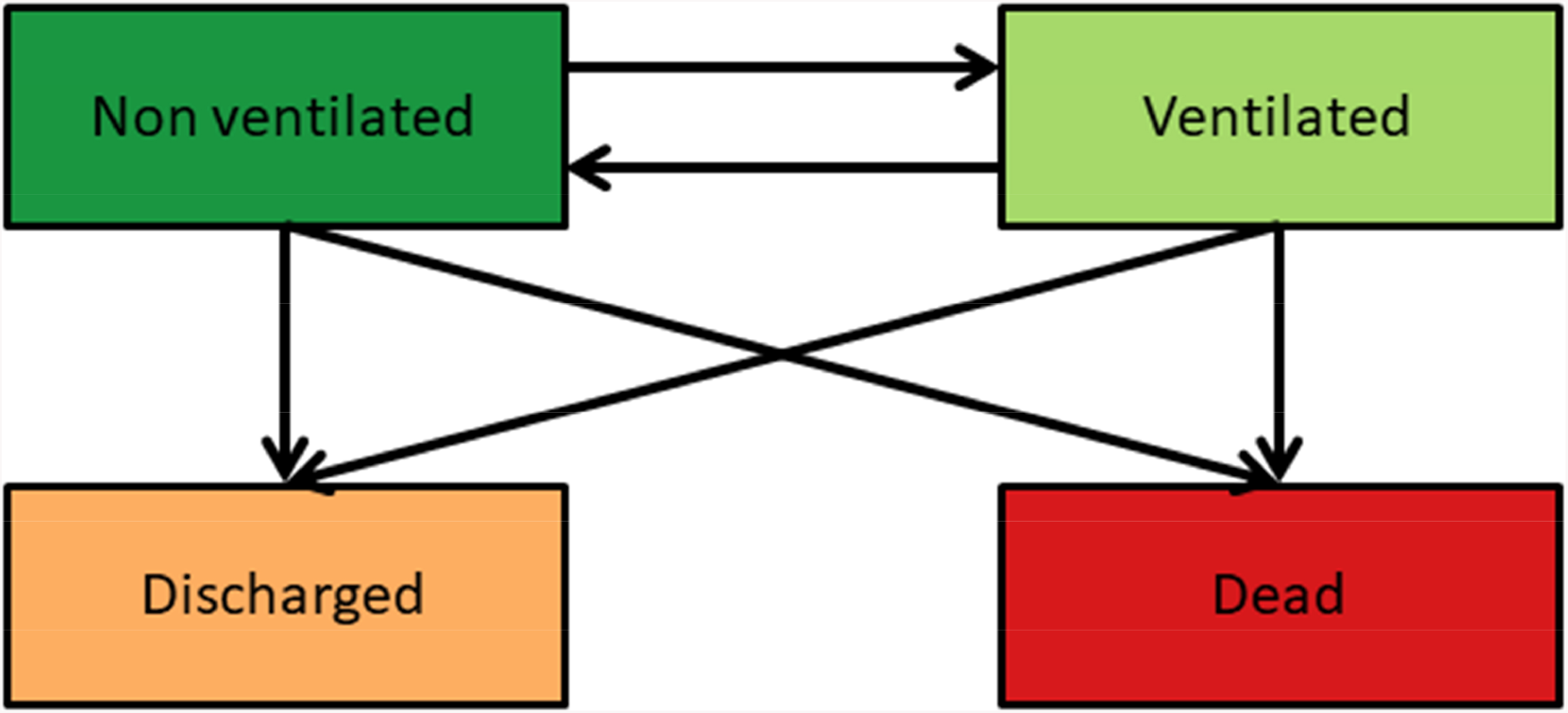
Multistate model for COVID-19 trials with a population of severe cases.

The course of a patients hospital stay in the treatment and control groups can be visualized with a stacked probability plot (11) (Figure 2). Important events of interest are visualized simultaneously over time in a single informative plot. The results of clinical trials are directly comparable if the stacked probability plots are published as a complement to the statistical analysis. Differing lengths of follow-up are accounted for by the time-dependent graphical display of the probabilities of being in one of the states of interest. Moreover, the events in this multistate model are objective and not subject to selection bias due to clinical judgement.

**Figure 2:**
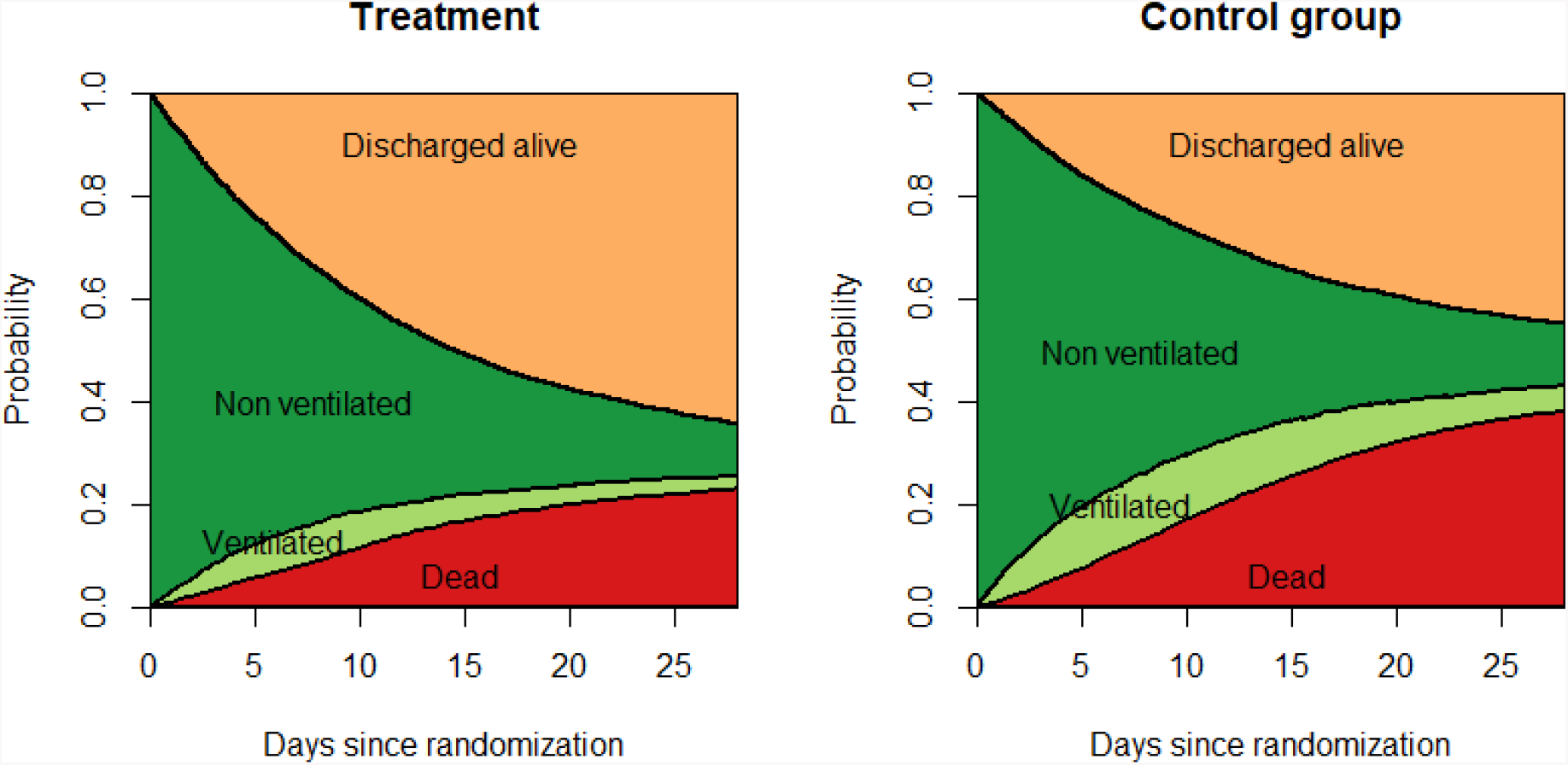
Stacked probability plot for the multistate model in Figure 1. The plot is based on a simulated data setting and illustrates for the treatment and respectively control group the probability to be mechanically ventilated, hospitalized without ventilation, discharged alive and dead over the course of time.

For the stacked probability plots the following information needs to be recorded:

– Hospital admission and discharge dates
– Vital state at end of follow-up
– Death date, if applicable
– Dates of mechanical ventilation

For clinical trials that have already been started, we highly recommend to provide the stacked probability plots. Specifically clinical trials using the categorical endpoints recommended by the WHO R&D Blueprint expert group have all the necessary information available for this detailed analysis.

For the planning of novel trials, we urgently suggest to analyze the available data with the multistate model. Etiology of treatment can be additionally studied with cause-specific Cox regression for all possible transitions in the model (12). Differences in duration of hospitalization and time spent under mechanical ventilation between treatment and control group can be modelled with the transition probabilities of the multistate model (13). The model avoids common pitfalls such as competing risks bias when studying hospital mortality and immortal time bias when considering mechanical ventilation (14).

A model that allows for more detailed information of treatment effect in mild and moderate cases is shown in Figure 3. Analysis of the model in Figure 3 is in principle the same as for the model in Figure 1. Treatment effects on being in one of the states at a particular time point can be visualized with the stacked probability plot and quantified with cause-specific hazard ratios for all possible transitions in the multistate model.

**Figure 3:**
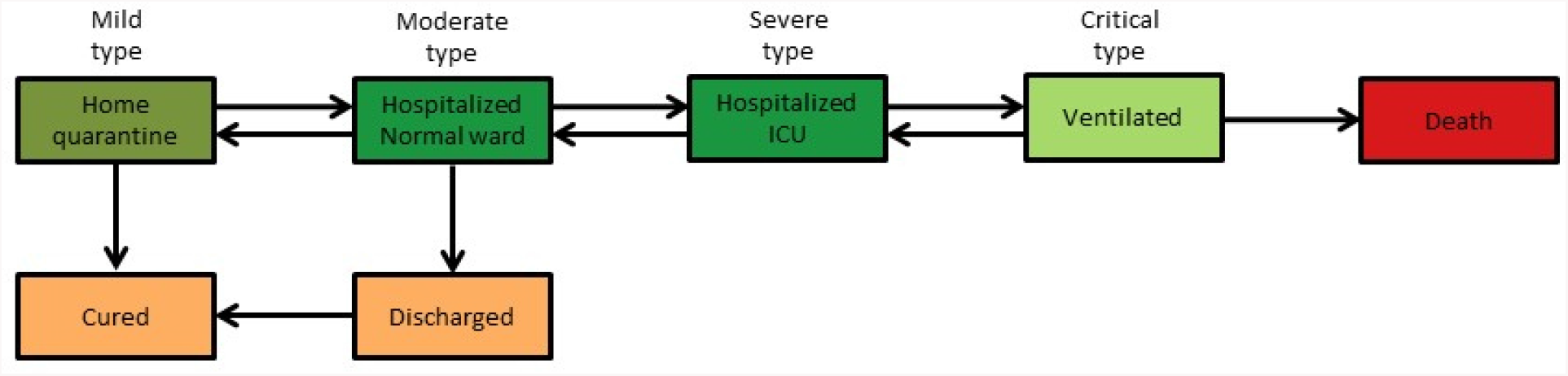
Detailed multistate model to allow for more sensitive endpoints in moderate and mild cases. The colors indicate how the model relates to the multistate in Figure 1. The discharge state in Figure 1 can be considered as a combination of “Discharged” and “Cured”. “Hospitalized Normal ward” and “Hospitalized ICU” describe in more detail the state “Non ventilated” in Figure 1. The labels “mild type” to “critical type” for the various states shall only give an idea on how the model accounts for the different types of severity of COVID-19. We do not claim to give a precise clinical definition.

Explanations on multistate model analysis and software are available in the literature (4,5,12,15–17).

## Discussion

Our review of registered interventional COVID-19 trials revealed high heterogeneity in the definition of primary endpoints. Most of the studies used a recovery endpoint for the primary analysis. We propose to accompany the primary analysis with multistate methodology. We highlight, that literature provides an extensive statistical tool box of suitable models to analyze clinical trials. All ongoing clinical trials, especially those with severe cases, should accommodate primary analysis with a stacked probability plot of the major events mechanical ventilation, discharge alive and death. The resulting informative plot is a powerful tool to harmonize the diversity of clinical endpoints and lengths of follow-up and thereby fastening accessibility of evidence and thus decision making. Additionally, treatment effects on a number of (potentially clinically opposite) endpoints can be studied simultaneously over time using cause-specific Cox regression and by estimating sojourn time spent in the various states. Moreover, while mechanical ventilation, hospitalization and mortality are objective outcomes, recovery and other states of severity are influenced by clinical judgement and may be additionally subject to measurement errors.

Besides these potentials, both of the proposed multistate models have the limitation that they do not account for patient triage possibly necessitated by ICU congestion (19). Additional research is needed to avoid selection bias arising from this special clinical situation. Finally, our proposal should be understood as a complement for the primary analysis. Rather than suggesting COS for COVID-19 trials, we provide a way for simultaneous evaluation of multiple endpoints within COS using information all of the available data.

To conclude, we recommend to complement the primary analysis with a stacked probability plot for the clinical events mechanical ventilation, discharge alive and death.

## Data Availability

not applicable

## Acknowledgment

We thank Dr. Klaus-Dieter Wolkewitz for valuable clinical input and Dr. Gerta Rücker and Dr. Erika Graf for reviewing the manuscript and providing important and helpful comments.

## Notes

### Competing Interest Statement

The authors have declared no competing interest.

### Funding Statement

MvC has been funded by the EQUIP Medical Scientists Programme of the University of Freiburg.

